# Haematology dimension reduction, a large scale application to regular care haematology data

**DOI:** 10.1101/2024.08.29.24312784

**Authors:** Huibert-Jan Joosse, Chontira Chumsaeng-Reijers, Albert Huisman, Imo E Hoefer, Wouter W van Solinge, Saskia Haitjema, Bram van Es

**Author notes:** Contributing authors.

## Abstract

**Background:** The routine diagnostic process increasingly entails the processing of high-volume and high-dimensional data. This processing may provide scaling issues that limit the implementation of these types of data into research as well as integrated diagnostics in routine care. Here, we investigate whether we can use existing dimension reduction techniques to provide visualisations and analyses for a complete bloodcount (CBC) while maintaining representativeness of the original data. We considered over 3 million CBC measurements encompassing over 70 parameters of cell frequency, size and complexity from the UMC Utrecht UPOD database. We evaluated PCA as an example of a linear dimension reduction techniques and UMAP, TriMap and PaCMAP as non-linear dimension reduction techniques. We assessed their technical performance using quality metrics for dimension reduction as well as biological representation by evaluating preservation of diurnal, age and sex patterns, cluster preservation and the identification of leukemia patients.

**Results:** We found that PCA performs systematically better than the UMAP, TriMap and PaCMAP in representing the underlying data. Biological relevance was retained for periodicity in the data. However, we also observed a decrease in predictive performance of the reduced data for both age and sex, as well as an overestimation of clusters within the reduced data. Finally, we were able to identify the diverging patterns for leukemia patients after use of dimensionality reduction methods.

**Conclusions:** We conclude that for hematology data, the use of unsupervised dimension reduction techniques should be limited to data visualization applications, as implementing them in diagnostic pipelines may lead to decreased quality of integrated diagnostics in routine care.

## 1 Introduction

The diagnostic process in a routine healthcare setting increasingly produces data in high volume, dimensionality and in multiple modalities, both structured and unstructured. Examples of these diagnostic data are ‘omics’ data such as transcriptomics, proteomics and metabolomics as well as imaging data, yet routine haemocytometer data of a complete blood count (CBC) can also be considered high-dimensional data. Visualisation of the data in a comprehensive way can be a challenge due to the high dimensionality. More importantly, to help healthcare professionals interpret these data for the benefit of individual patients, integration of the different types of data into integrated diagnostics models is warranted. One of the modelling challenges in the development and deployment of these models is the combination of vast data volumes and their high dimensionality, which may lead to computational performance issues. There is thus a need to ensure feasibility of integrated diagnostics models. One of the ways to achieve this is by using a low-dimensional representation of these data rather than the full dataset. Such a representation can be generated using dimension reduction techniques.

Dimension reduction has historically been performed by the use of principal component analysis (PCA). This linear transformation technique assumes normally distributed variables, and is primarily focused on establishing a dimension reduction that is preserving the global structure. Global structure preservation aims at preserving the global patterns in the data, such as obvious clusters that are present in the data, whereas the local structure preservation aims at preserving more intrinsic patterns in the data, i.e. preserving the neighbourhood for each point. Several more recent dimension reduction approaches aim to also preserve local structure. One way to do this is through (non-linear) manifold approximation, which is based on learning the underlying structure of the data, mostly based on nearest neighbours. Some examples for these type of methods are Uniform Manifold Approximation and Projection (UMAP)[1], Pairwise Controlled Manifold Approximation (PaCMAP)[2], and TriMap, a triplet-based approach [3].

Applying these methods to high-dimensional biological data has been performed before, including flow cytometry workflows, transcriptomics data, RNA sequencing data, and protein structure analysis among others [4, 5, 6, 7, 8, 9]. However, to the best of our knowledge, comparative work on robustness of dimension reduction on large haematological data has not been performed before.

A complete blood count (CBC) assessing red and white blood cells and platelets, is one of the most frequently performed diagnostic procedures. Haemocytometers, on the basis of flow-cytometry, use proprietary algorithms to combine cell characteristics such as size, granularity, lobularity and viability into clinically relevant parameters like hemoglobin levels or white blood cell differentiation patterns. However, next to these parameters currently reported to the clinic, each routine haematology measurement actually encompasses research-only values and raw cell characteristics of red and white blood cells and platelets that are currently not used in clinical care. In the University Medical Center Utrecht (UMCU), Utrecht, the Netherlands, the raw hematology data of over 3 million samples that were measured on Abbott CELL-DYN Sapphire hematology analyzers were stored in the Utrecht Patient Oriented Database (UPOD) since 2005. The full content and extent of the database is described elsewhere [10]. Previous UPOD research shows there is biologically and clinically relevant information hidden in the unreported hematology measurements of these samples [11, 12, 13, 14, 15, 16]. Using dimension reduction methods to enable processing of raw CBC and visualising or combining it into integrated diagnostics models may therefore eventually improve clinical practice.

Considering this vast amount of haematological data, and its high number of dimensions, we set out to find a robust approach in reducing the dimensions of the data, so that it can be better processed but also better visualized. By investigating the performance of the dimension reduction methods, we aim to ensure their usability in routine haematological data to improve clinical care, for example in diagnostic pipelines. As a dimension reduction should be a good representation of the original data, we not only compared several current dimension reduction techniques (PCA, UMAP, TriMap, and PaCMAP) in their ability to both capture global and local structure, but also assessed their ability of preserving biological relevancy of the data.

## 2 Results

### 2.1 Descriptives

In total, we extracted 3, 093, 792 samples from 358, 614 unique patients. 52.8% of the samples were from male patients, the median age at measurement was 51 (IQR: 27-66). After preprocessing the haematological data, we applied imputation to 1, 107, 049 samples for haematological parameters that were missing as a result of laboratory protocols (e.g., not using reticulocyte mode). Preprocessing of the data included the clipping of haematological parameters, to mitigate distorting the dimension reduction due to extreme outliers. The clippings of the parameters are given in supplemental table S1.

### 2.2 Dimension Reductions

#### 2.2.1 Parameter Tuning Results

##### Number of Neighbours

First, we compared the amount of nearest neighbours used for dimension reduction for both UMAP, TriMap, and PaCMAP. The results are shown in figure 1 for UMAP, and in figure S1 for PacMAP. For an increasing sample size and number of neighbours, we observed an initial improvement with a rapid stagnation (1, figure S1).

**Fig. 1:**
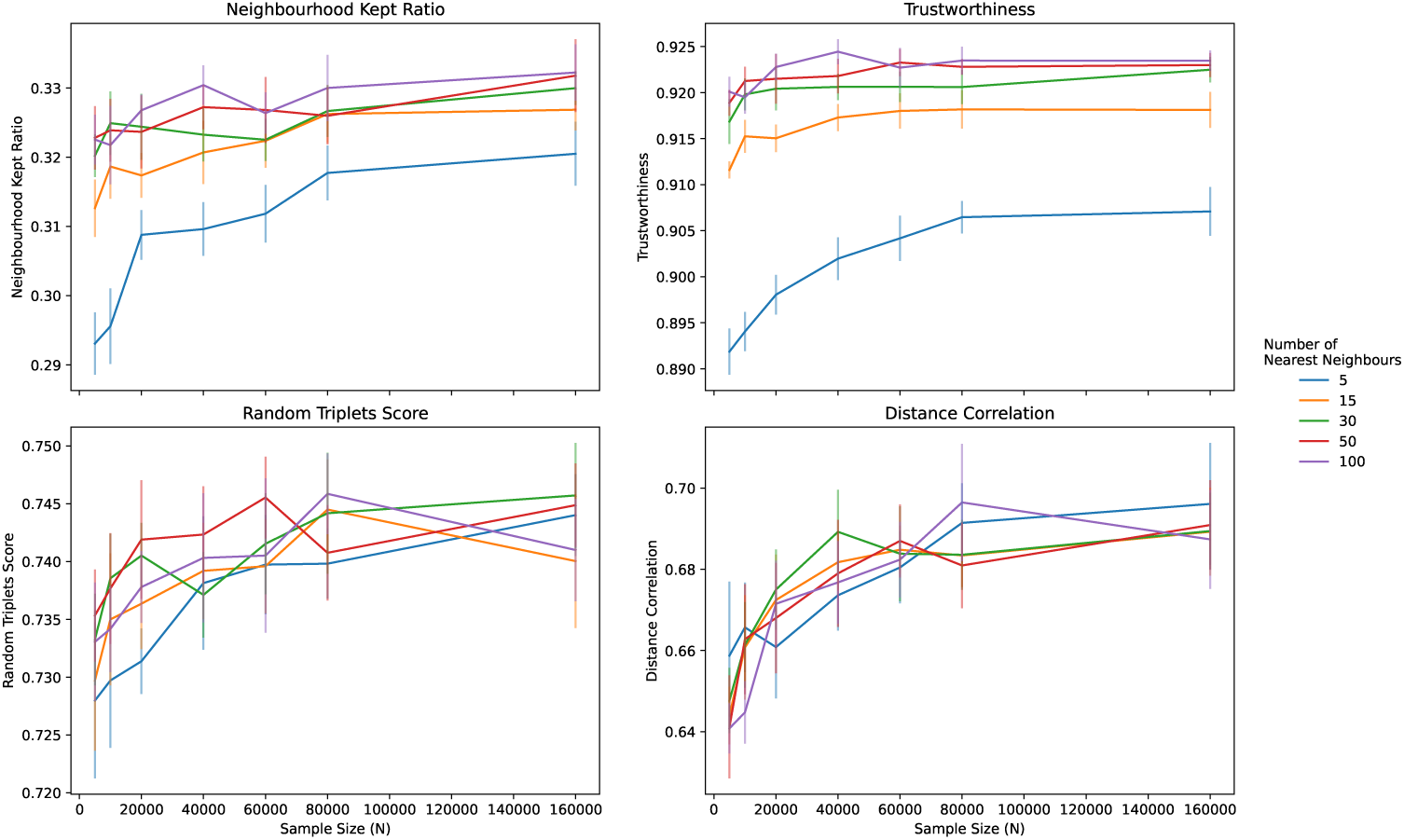
Evaluating UMAP with quality metrics across different numbers of neighbours and sample size along with the 95% Standard Error (SE) for each sample size and number of nearest neighbours

The neighbourhood kept ratio ranged from 0.29 at 5000 samples and 5 nearest neighbours to 0.33 at 160,000 samples and 100 nearest neighbours for UMAP. However, the scores for any number of neighbours from 15 and above were similar with increasing sample size. For trustworthiness, using 5 nearest neighbours yielded worse results than using 15 or more neighbours (0.89-0.91). For the other number of neighbours, the performance was limited to 0.92. For global distance preservation methods, UMAP was stable for random triplet score ranging from 0.72 for 5 nearest neighbours at 5000 samples to 0.74 for all number of neighbours at 40,000 to 160,000 samples. Distance correlation increased from 0.64 at 5000 samples to 0.69 at 40,000 to 160,000 samples (figure S2).

For PaCMAP, a similar pattern was observed (figure S1). Local distance preservation as measured through the neighbourhood kept ratio ranged from 0.33 at 5000 samples and 5 nearest neighbours to 0.36 at 160,000 samples with 30, 50, or 100 nearest neighbours. Trustworthiness ranged from 0.88 at 5000 samples for 5 nearest neighbours to 0.90 at 160,000 samples for all number of nearest neighbours. However, this performance was already reached at 10,000 samples by using 30, 50, or 100 nearest neighbours. Global distance preservation as measured by the random triplets score ranged from 0.73 to 0.74 for all number of neighbours. Distance correlation remained relatively stable, with scores ranging from 0.66 to 0.67.

Considering the results, we decided to limit the sample sizes to 40,000 for TriMap to find the number of inand outlying neighbours, because increasing the sample beyond this point yielded similar result, yet dramatically increased computational costs (data not shown). The result of this tuning are found in figure S2. We observed no large differences for the number of outliers used for TriMap. However, we did observe substantial increased for global distance preservation metrics when increasing the number of inliers. Random triplets score increased from 0.75 (5 inliers) to 0.78 (100 inliers). Distance Correlation increased from 0.75 (5 inliers) to 0.81 (100 inliers). However, we decided to move forward with 50 inliers and 15 outliers for TriMap, since increasing the number of neighbours was computationally not feasible for the entire data set of over 3 million samples (data not shown).

##### Number of Components

As we used 50 nearest neighbours for the dimension reductions, we also used 50 nearest neighbours for calculating the dimension reduction quality metrics. We then increased the number of components for the final dimension reduction. We used 40,000 random samples, which were matched across the dimension reduction methods.

We compared 2, 4, 6, 8, 10, 20 and 30 components to get a rough estimation of the increase in performance for each of these models. Figure 2 shows the results for the neighbourhood kept ratio, trustworthiness, random triplet score and distance correlation. For all scores, PCA performed best across all number of component (p*<* 0.001) (figure 2). Additionally, performances for UMAP, TriMap, and PaCMAP barely increased by increasing the number of components.

**Fig. 2:**
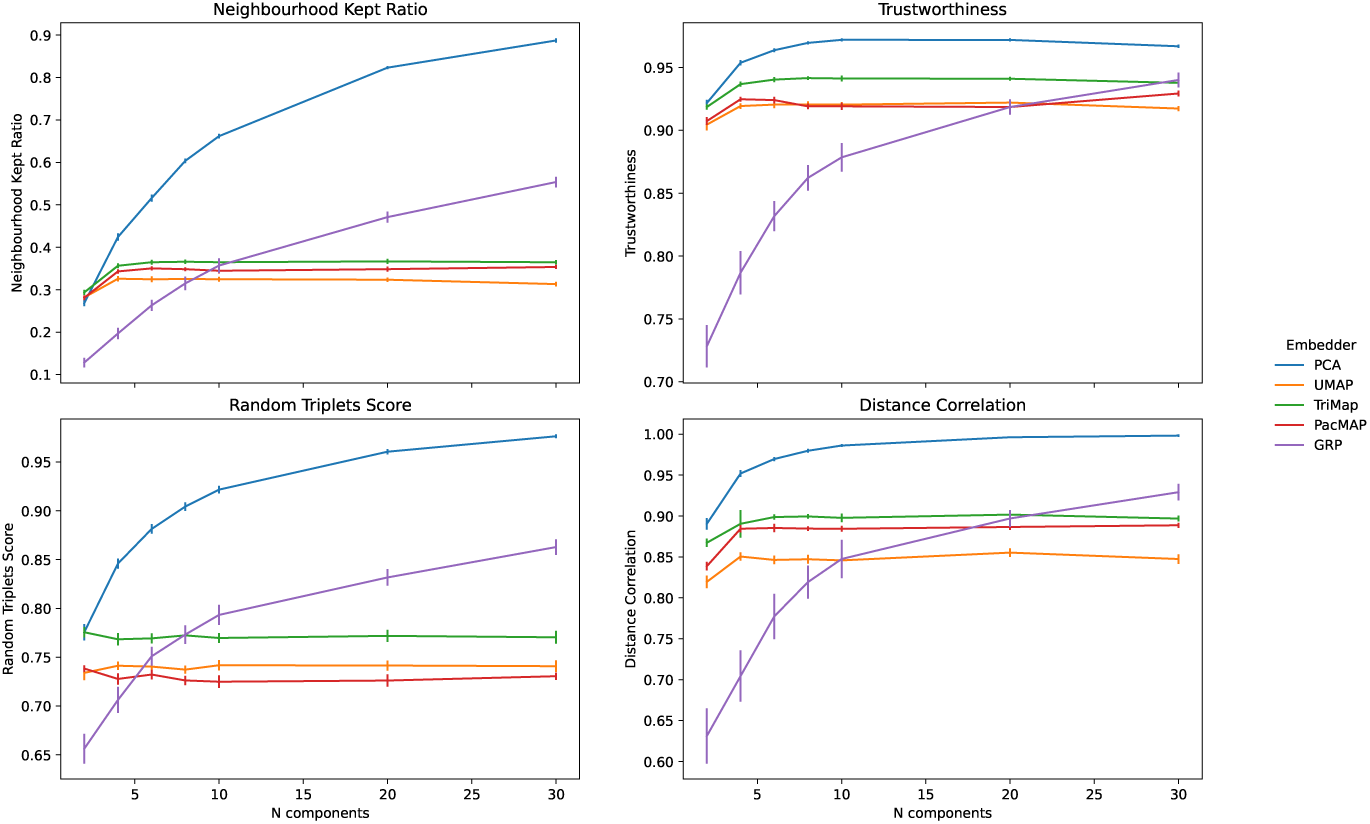
Dimension reduction metrics across a different number of dimensions along with the 95% SE for each number of components

Focussing on local distances, the neighbourhood kept ratio increased from 0.27 (2 dimensions) to 0.89 (30 dimensions) for PCA, whereas it stagnated for UMAP around 0.32, TriMap around 0.36, and PaCMAP around 0.35. GRP increased from 0.13 (2 dimensions) to 0.55 (30 dimensions). Trustworthiness was high for all dimension reduction methods, except for GRP at lower dimensions. PCA (0.92-0.97) had the highest scores, UMAP and PaCMAP performed similarly (0.90-0.92 for UMAP; 0.91-0.93 for PaCMAP). TriMap performed better than the other manifold approaches (0.92–0.94). GRP performed worse at lower dimensions (ranging from 0.73 to 0.94).

When it comes to global distances, PCA outperformed all other dimension reduction methods on both random triplets score (0.78 to 0.98) and distance correlation (0.81 to 0.91, max = 0.93 at 8 dimensions). The random triplets score remained stable for the three manifold approaches, scoring 0.74 for UMAP, 0.78 for TriMap, and 0.73 for PaCMAP. GRP increased from 0.66 at 2 dimensions to 0.86 at 30 dimensions. Distance correlation for the manifold approaches increased primarily with lower dimensions, scoring 0.90 for UMAP, 0.91 for PaCMAP and 0.92 for Trimap to 0.92 for UMAP, 0.93 for PaCMAP and 0.94 for TriMap at 4 components, which then remained stable with increasing dimensions.

Although we observed an increase of performance for PCA and GRP with increasing components, we also observed a stagnation for manifold approaches at 4 components. Considering the increasing computational complexity of the manifold approaches with increasing components, we decided to limit the number of components to 6 for all methods, in order to reduce the entire data set of over 3 million samples.

### 2.3 Preservation of Biological Representation

#### 2.3.1 Explorative analysis

One of the broad epistemic features of, at least part of, the hematology parameters is the presence of a diurnal pattern[17, 18]. We should expect that dimension reduction algorithms preserve such broad qualitative features. Here we consider the diurnal patterns of the reduced dimensions, including a cosine-fit.

Figure 3 shows the 6 UMAP dimensions. We observed a diurnal pattern for each of the components, primarily split between daycare (6:00–18:00) and care during the night, with clear progression within the daycare time-period. The clearest diurnal patterns in the non-reduced data are obtained for the neutrophil and the eosinophil fractions. For these fractions, and all components for the dimension reduction techniques, we observed significant results for the cosine-fit (table 1). Additionally, figure 4 shows the retention of periodicity in the dimension reductions compared to the periodicity of the neutrophil fraction. We chose neutrophil fraction as this parameter has a clear diurnal evolution.

**Fig. 3:**
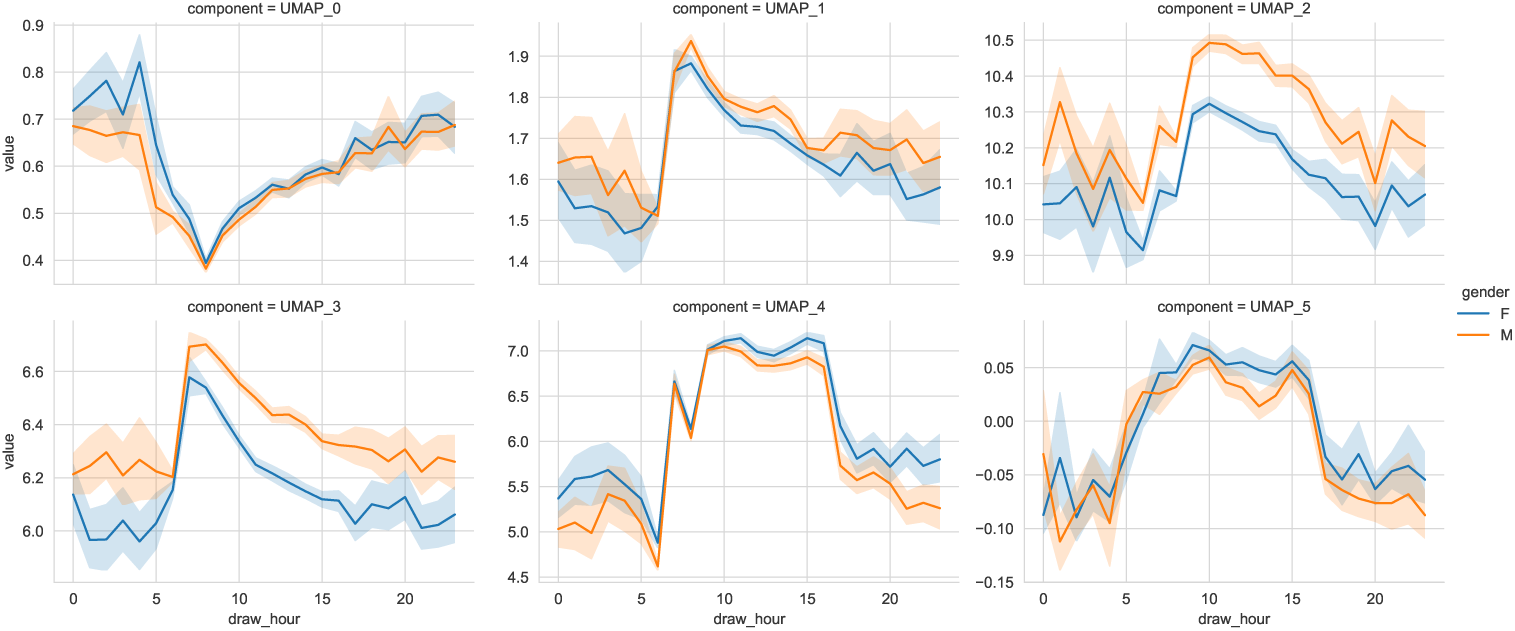
Intraday variation of UMAP dimensions showing a clear diurnal pattern, as expected from prior work showing diurnality of hematology parameters, as well an offset between men/women in the 3*^rd^* and 4*^th^* dimensions

**Table 1:** Results of a Cosine-fit. The dimension reductions for this result were extracted from a sample of 100.000 measurements, repeated 10 times.

| Reduction technique | component | amplitude | offset | bias | p-value |
| --- | --- | --- | --- | --- | --- |
| PCA | 0 | $3.26 \pm 0.051$ | $0.80 \pm 1.44$ | $0.67 \pm 1.55$ | $1e-16$ |
| | 1 | $1.00 \pm 0.016$ | $-1.85 \pm 0.033$ | $-0.43 \pm 0.012$ | $1e-16$ |
| | 2 | $1.07 \pm 0.041$ | $-0.80 \pm 0.021$ | $0.20 \pm 0.019$ | $1e-16$ |
| | 3 | $1.57 \pm 0.037$ | $-0.26 \pm 0.017$ | $0.73 \pm 0.018$ | $1e-16$ |
| | 4 | $0.40 \pm 0.009$ | $-1.61 \pm 0.040$ | $-0.12 \pm 0.009$ | $1e-16$ |
| | 5 | $0.82 \pm 0.011$ | $0.56 \pm 0.019$ | $0.52 \pm 0.006$ | $1e-16$ |
| GRP | 0 | $2.01 \pm 1.15$ | $-1.08 \pm 1.94$ | $0.11 \pm 1.31$ | $1e-16$ |
| | 1 | $1.44 \pm 1.23$ | $-0.35 \pm 1.52$ | $0.21 \pm 0.82$ | $1e-16$ |
| | 2 | $1.70 \pm 0.84$ | $0.27 \pm 1.61$ | $0.38 \pm 0.96$ | $1e-16$ |
| | 3 | $1.51 \pm 0.78$ | $1.33 \pm 1.75$ | $0.0046 \pm 0.90$ | $1e-16$ |
| | 4 | $1.02 \pm 0.54$ | $0.81 \pm 1.76$ | $-0.058 \pm 0.55$ | $1e-16$ |
| | 5 | $1.44 \pm 0.10$ | $1.28 \pm 1.88$ | $-0.37 \pm 0.73$ | $1.5e-3$ |
| UMAP | 0 | $0.13 \pm 0.096$ | $0.48 \pm 2.04$ | $2.24 \pm 3.67$ | $1e-16$ |
| | 1 | $0.15 \pm 0.043$ | $-1.14 \pm 2.18$ | $1.24 \pm 1.50$ | $1e-16$ |
| | 2 | $0.19 \pm 0.070$ | $1.69 \pm 1.92$ | $9.31 \pm 1.86$ | $1e-16$ |
| | 3 | $0.87 \pm 0.510$ | $-0.95 \pm 0.85$ | $5.20 \pm 1.45$ | $1e-16$ |
| | 4 | $0.70 \pm 0.509$ | $1.31 \pm 1.42$ | $4.82 \pm 1.54$ | $1e-16$ |
| | 5 | $0.11 \pm 0.059$ | $-1.20 \pm 1.30$ | $1.17 \pm 2.05$ | $1e-13$ |
| TriMap | 0 | $15.82 \pm 0.24$ | $0.74 \pm 1.44$ | $3.55 \pm 8.1$ | $1e-16$ |
| | 1 | $5.10 \pm 0.06$ | $-1.21 \pm 0.024$ | $2.26 \pm 0.125$ | $1e-16$ |
| | 2 | $5.05 \pm 0.20$ | $-0.61 \pm 0.014$ | $2.86 \pm 0.133$ | $1e-16$ |
| | 3 | $7.94 \pm 0.19$ | $-0.38 \pm 0.010$ | $5.42 \pm 0.147$ | $1e-16$ |
| | 4 | $1.40 \pm 0.08$ | $-0.98 \pm 0.059$ | $0.52 \pm 0.069$ | $1e-16$ |
| | 5 | $0.73 \pm 0.053$ | $0.32 \pm 0.060$ | $0.54 \pm 0.029$ | $1e-16$ |
| PaCMAP | 0 | $2.15 \pm 0.033$ | $0.71 \pm 1.440$ | $0.43 \pm 0.99$ | $1e-16$ |
| | 1 | $0.54 \pm 0.018$ | $-1.19 \pm 0.040$ | $-0.069 \pm 0.015$ | $1e-16$ |
| | 2 | $0.82 \pm 0.024$ | $2.65 \pm 0.025$ | $-0.35 \pm 0.01$ | $1e-16$ |
| | 3 | $0.99 \pm 0.020$ | $2.85 \pm 0.017$ | $-0.50 \pm 0.015$ | $1e-16$ |
| | 4 | $0.38 \pm 0.030$ | $2.26 \pm 0.075$ | $-0.081 \pm 0.020$ | $1e-16$ |
| | 5 | $0.19 \pm 0.041$ | $-1.72 \pm 0.090$ | $-0.047 \pm 0.016$ | $1e-16$ |
| Neutrophil fraction | n/a | $7.3 \pm 0.1$ | $0.1 \pm 0.01$ | $68.9 \pm 0.08$ | $1e-16$ |
| Eosinophil fraction | n/a | $0.24 \pm 0.002$ | $-2.8 \pm 0.01$ | $0.85 \pm 0.002$ | $1e-16$ |

**Fig. 4:**
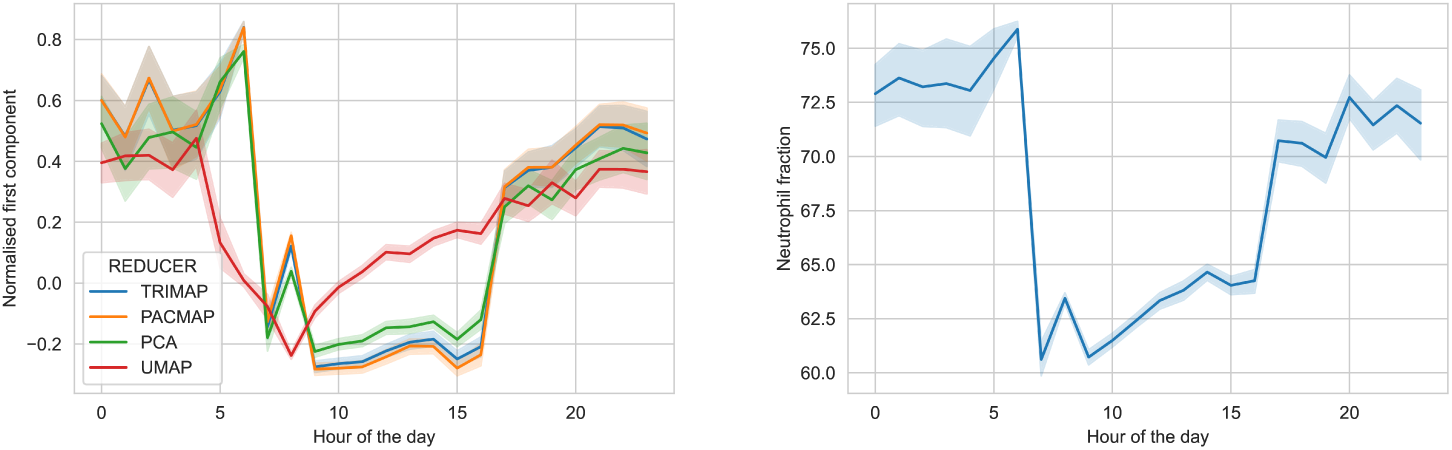
Time evolution of (left) first component for different reducers with 95% SE, (right) the neutrophil fraction.

#### 2.3.2 Prediction performance

To assess preservation of biological relevance, we compared age (*≤* 20 versus *≥* 60) and sex prediction performance of original data to the prediction performance of reduced data. Results of age at sampling predictions can be found in figure 5. We used data from 170,000 random samples and matching the samples to their reduced data, and used 30,000 random samples for validation. We observed a significant (p *<* 0.001) drop in performance when data from any dimension reduction method was used. We observed very stable performances across the 10-fold cross validation, resulting in small variation for the accuracy and MCC. While the original data showed higher performance (accuracy = 0.88, MCC= 0.74) for age-classification, we observed a lower accuracy, ranging from 0.76 for GRP to 0.80 for the manifold methods (PCA = 0.79, and a lower MCC, ranging from 0.47 for GRP to 0.56 for TriMap (PCA = 0.55; UMAP = 0.55; PaCMAP = 0.56). This meant that applying dimension reduction negatively impacted classification tasks. The same pattern was observed in sex prediction (figure S5). The original data showed an accurary of 0.76 and a MCC of 0.51. For the data in reduced space, the accuracy ranged from 0.61 for GRP to 0.7 for UMAP and TriMap (PCA = 0.68; PaCMAP = 0.69. The MCC ranged from 0.18 for GRP to 0.39 for UMAP (PCA = 0.34; TriMap = 0.38; PaCMAP = 0.36).

**Fig. 5:**
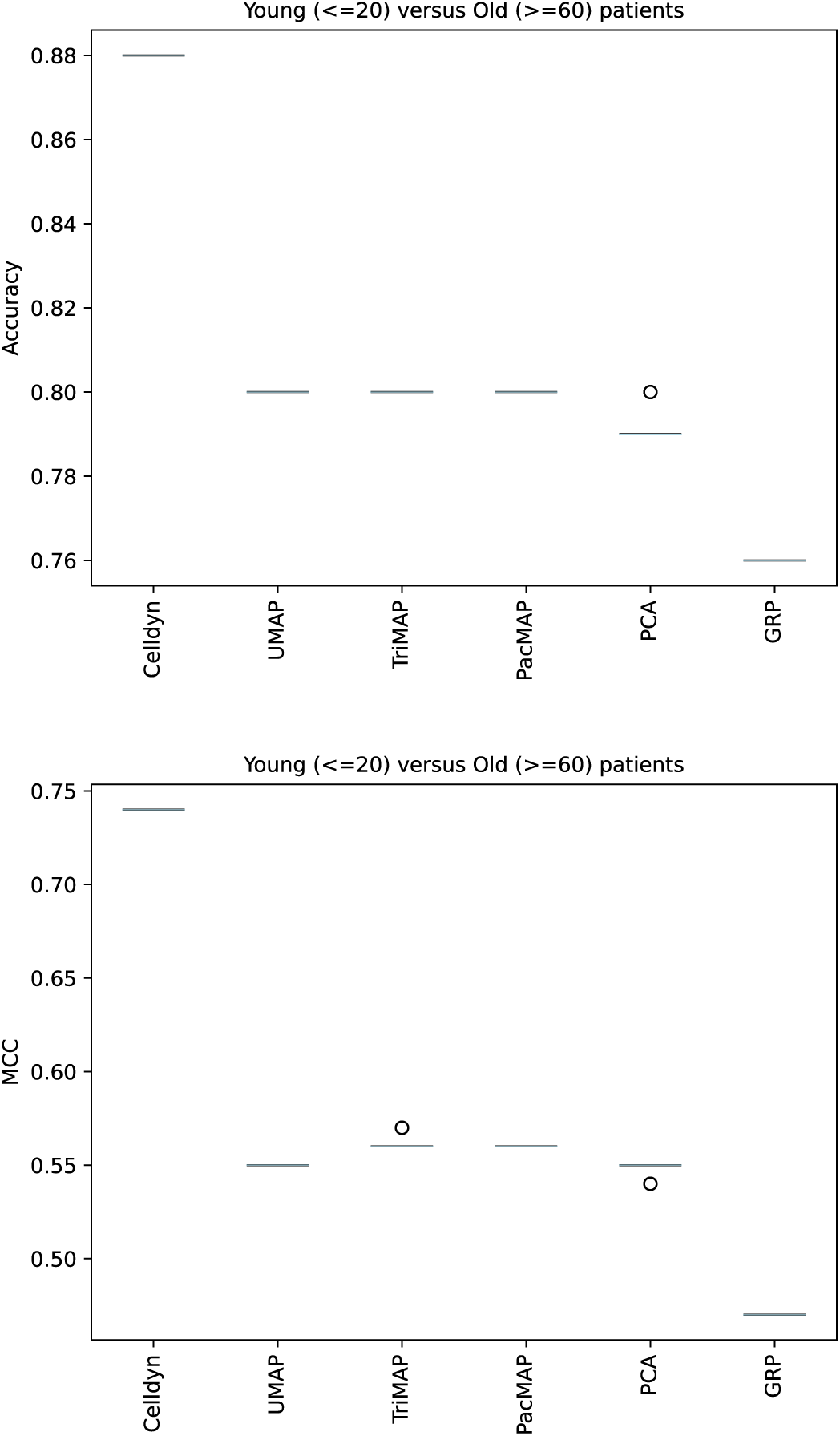
Predictive performance for patients *<* 20 (’young’) versus patients *>* 60 (’old’) in the dedicated validation set. The prediction performances dropped significantly (p *<* 0.001) after applying dimension reduction techniques. *n* = 170,000 for training; *n* = 30,000 for validation; *n folds* = 10.

#### 2.3.3 Cluster Preservation

Table 2 shows the performances of clustering methods using the reduced data. We observed an excess of clusters with subsequent low values for the Normalised Mutual Information (NMI) score and Adjusted Rand Index (ARI), showing that the dimension reduction methods have a tendency to generate an excess of clusters in comparison with the real data. We identified 12 clusters in the original data, whereas we found 32, 31 and 12 for PCA at 3, 6 and 12 components respectively. For the manifold approaches, we found a large inflation of clusters. For UMAP we identified 115, 84, and 81 clusters; for TriMap we identified 45, 44 and 53 clusters; for PaCMAP we identified 42, 43 and 54 clusters, all with 3, 6 and 12 components respectively. Finally, for GRP we identified 30, 22 and 5 clusters for 3,6 and 12 components respectively.

**Table 2:**
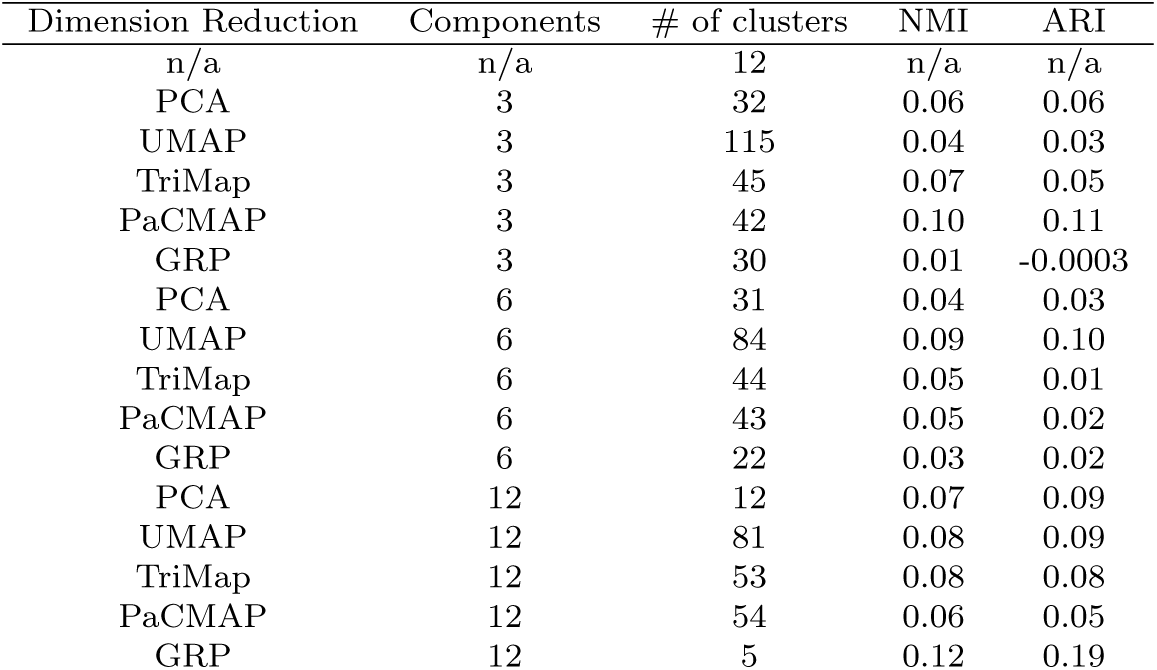
Comparison of cluster alignments, using a pipeline with a standard scaler, a dimension reduction method and a clustering model. As a default we used the Manhattan distance, with 50 neighbours and a random selection of 100.000 samples from the haematology set. Relevant hardware information: Xeon W-2125 at 4GHz and 8 logic cores with 64GB memory

Comparing the NMI score and ARI we found that, overall, scores were low (*≤* 0.10 for NMI and ARI, and did not improve when increasing the number of components, except for GRP with a NMI of 0.01 at 3 components and 0.12 at 12 components, and an ARI of −0.0003 at 3 components to 0.19 (table 2).

Furthermore, we found that, in terms of cluster quality, UMAP stagnates to a value well under the optimum for increasing number of components being on par with PCA for smaller number of components (figure S6). Additionally, we observed that all manifold approaches maintain a high level of cluster-inflation for increasing number of reduced dimensions. Finally, we observed that for a low number of reduced dimensions, all tested dimension reduction techniques produced a considerable inflated number of clusters as detected by HDBSCAN compared to the baseline cluster detection on the original data (fig 6).

**Fig. 6:**
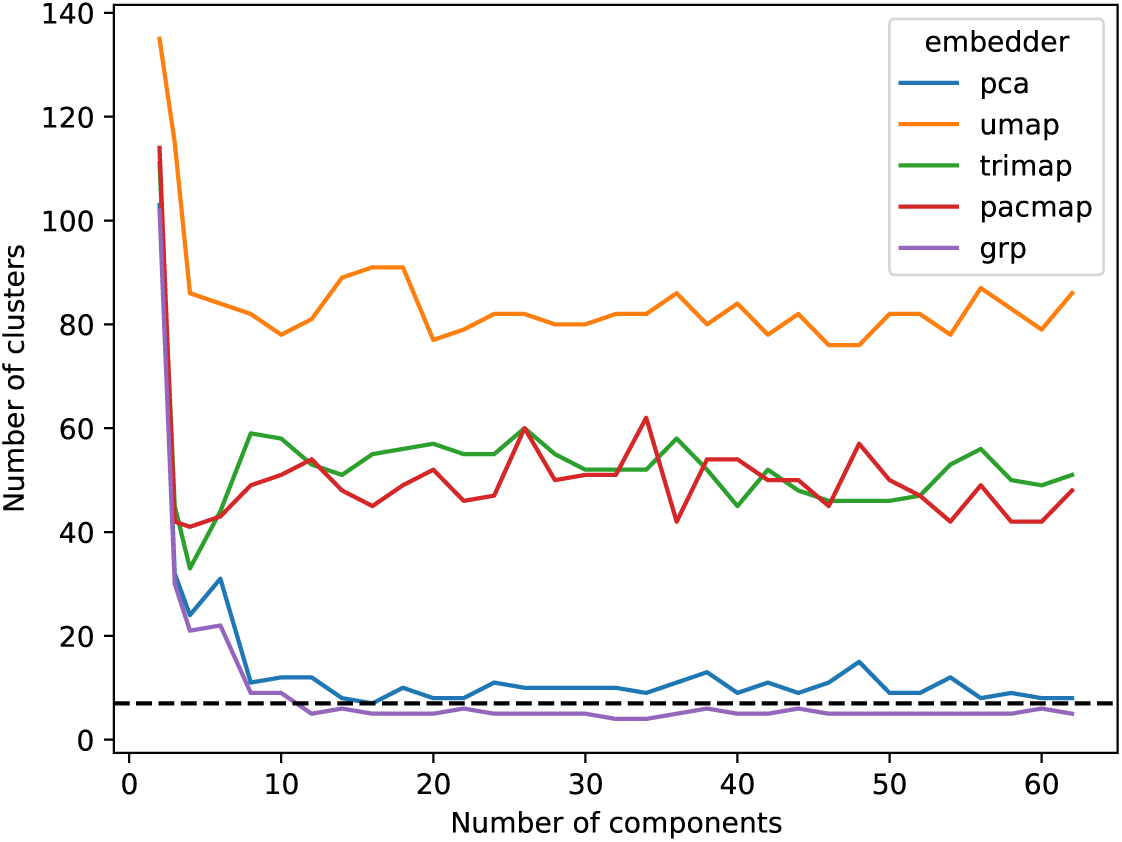
Number of clusters extracted using HDBSCAN. Here, dimension reduction was performed with PCA, UMAP, TriMap, PaCMAP, or GRP on 10.000 samples.

#### 2.3.4 Identification of Leukemia-Like Patients

In the original data (figure 7) we found significant differences between patients that were identified as having chronic lymphatic leukemia (CLL) with respect to our overall population for both white blood cell count as lymphocyte count. In total, we identified 3205 samples from patients with CLL, and compared these samples to all other samples in the data (*n* = 3,090,580). For all dimension reductions, we found similar results, where the CLL patients’ data had significantly different distributions (p*<* 0.001) compared to the general population for a large portion of the dimensions (figures S7 to S11).

**Fig. 7:**
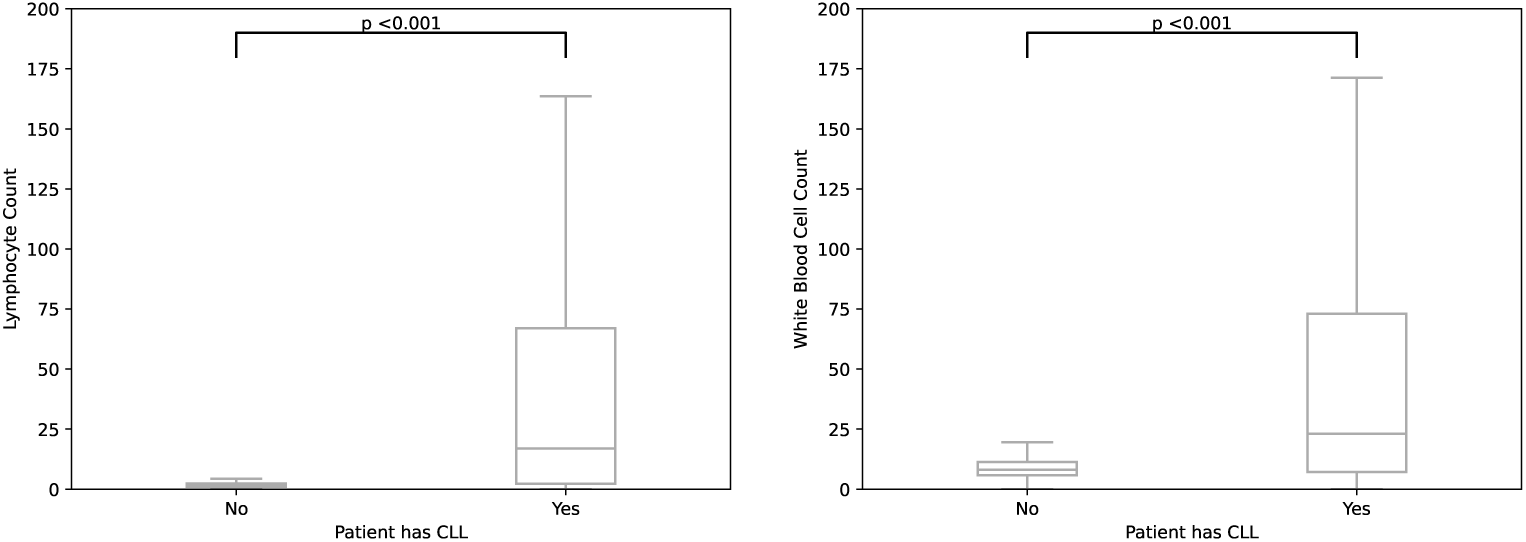
Boxplots showing the median and interquartile range of lymphocyte and leukocyte counts (x10^9^) of patients with CLL (*n* = 3, 205) versus patients without leukaemia (*n* = 3, 090, 580) in the original data

## 3 Discussion

In this study, we investigated the use of dimension reduction methods in a large set of routine CBC data from the Abbott CELL-DYN Sapphire haemocytometer. We compared PCA, UMAP, TriMap, and PaCMAP with multiple performance metrics (neighbourhood kept ratio, trustworthiness, random triplet score, and distance correlation). We found that looking at dimension reduction metrics, PCA was best performing in comparison with UMAP, TriMap and PaCMAP. As the purpose of these dimension reductions lie in analysis and interpretation, we investigated if any biological representation was correctly maintained. We found that diurnal patterns were maintained, but that predictive tasks (such as age and sex) performed significantly worse compared to the original data, and that clustering tasks resulted in an overestimation of clusters compared to the original data. We conclude that using dimension reductions will result in a loss of information compared to the original data, even in predictive tasks where subgroups should be apparently clear.

In literature, UMAP and other (non-linear) dimensionality reduction techniques are evaluated as superior with respect to PCA [19, 20, 2, 3]. However, the utility of UMAP and other nearest-neighbours-based dimension reduction methods is seemingly limited to very-low dimensional representations for the purpose of visualisation[2, 3]. In our study, we observe that for increasing dimension reduction dimensionality, the manifold techniques converge to dimension reduction scores that are far from optimal, whereas PCA reaches near-optimal scores well before it is able to explain 95% of the variance (n components = 30). This is likely the case for other global methods, but further research is needed to study this. We deem that this effect is partly because of the large sample size, meaning that the neighbourhood for a certain sample is harder to define, or we need a larger number of neighbours when increasing sample size. However, increasing the number of neighbours can result in computational issues, considering the pairwise nature of the dimension reduction techniques and performance measures.

When dealing with neighbourhood-based dimension reduction methods such as UMAP, TriMap and PaCMAP there is a trade-off between the preservation of local and global characteristics. Possible mitigations are to increase the number of components[21] and the number of nearest neighbours. The number of dimensions and neighbours is dependent on the amount of samples in the dataset. However, increasing the number of dimensions and number of neighbours increases the complexity of dimension reduction. Furthermore, for an increasing number of samples of multiple modalities, the heterogeneity of the data can increase, and it then becomes more difficult to embed the data with sufficient accuracy i.e., more samples does not inherently equate to a better dimension reduction. Finally, PCA becomes competitive in terms of dimension reduction performance when increasing the number of dimensions since the amount of explained variance increases, while being orders of magnitude more efficient computationally, especially if one considers the availability of Incremental PCA that has a constant memory complexity[22].

Another way to mitigate the issue with the trade-off between global and local characteristics, is to limit the number of samples used for the dimension reduction such that it contains enough samples per stratification, but not more. This requires enough information for the stratifications we are interested in, which in turn requires labelling. This is a known issue when using (routine) healthcare data, as the administrative start of a disease that is indicated by registration of a certain diagnosis does not coincide with the physical start of the disease. As the physical start of the disease may affect some or all parts of the CBC, labelling of disease presence at the time of blood draw is intrinsically difficult. Moreover, most patients that visit our tertiary care centre suffer from complex diseases and multiple comorbidities, further complicating labelling of our haematology data. Because of these issues, we were unable to retrieve clear labels for our samples.

One other mitigation of the problem with large sample sizes and neighbourhood-based dimension reduction methods that leads to improved tractability is the use of dimension reduction alignment, where we partition the datasets to create many dimension reductions that are subsequently aligned, using e.g. Procustes transformation [23]. Another benefit of dimension reduction alignment is that adding new data to the dimension reduction is much faster.

### 3.1 Biological performance

We investigated patterns in the data that are known to be present within haematology data. Indeed, we observed known diurnal patterns of white blood cells [24, 25, 18]. This pattern was also observed within the data after dimension reduction, showing preservation of intraday variation by the dimension reduction methods.

With respect to the prediction of samples belonging to subgroups in the data, we observed a significant decreased performance in the reduced data. We deem that dimension reduction before prediction tasks in these data is not a preferable approach, since the loss of information or quality of data representation is an apparent issue. Increasing the number of dimensions might mitigate this[21], but can lead to more complex dimension reduction processes, and we observed that the manifold approaches did not convergence to an optimal data representation of increasing dimensionality (figure 2). Rather than using dimension reduction, more emphasis should be given towards proper feature selection for analysis when the amount of parameters is too high for the amount of samples. This can, of course, be combined with dimension reduction [26]. In literature, we found some beneficial results of using dimension reduction before prediction in different settings, since it can offer similar or better model performance to using original data at least in experimental circumstances [27, 28, 29], or that dimension reduction can be used for feature selection [30]. However, this requires a robust dimension reduction method, which also preserves the distances when applying this method. Considering our findings, the use of unsupervised dimension reduction techniques before modelling should be approached with caution or even refrained from.

Finally, we applied clustering to assess the cluster preservation in the reduced data. We find that using dimension reduction will result in an overestimation of the amount of clusters when using HDBScan. This comes together with the loss of information, or quality. As mentioned before, this might be mitigable, but will increase computational costs and complexity. We do, however, observe that leukemia-like patients are indeed outliers after applying dimension reduction methods.

### 3.2 Further Research

Considering we have limited towards unsupervised non-parametric dimension reduction methods, a logical next step is to use supervised and/or parametric dimension reduction. An improvement of non-parametric UMAP is parametric UMAP where a learnable parameterised model sits between the dimension reductions and the final loss, enabling the addition of e.g. a global loss contribution [31]. Additionally, when we are dealing with large volume data, benefit might be gained from using a fully parameterised dimension reduction methods such as Differentiating dimension reduction Networks (DEN), which is more interpretable compared to UMAP and t-SNE because of the parametric nature [32]. Finally, when it comes to generalizability of dimension reduction results, and working towards a more holistic integrative approach of data analysis within healthcare, fully parameterised models such as variational autoencoders with contrastive-loss optimization are interesting from the perspective of transfer learning, as it adds flexibility to continue learning with incoming data and transferring the resulting model to other institutions to continue training on their on-premise data, which can play a role in federated learning. In addition, research could study the use of (semi-)supervised dimension reduction approaches. To ensure clinical relevance, sparsely available labels can be employed, and consequently semi-supervised UMAP/t-SNE or Multi-Class, Multi-Label (MCML) dimension reduction can be deployed. Possible other variables that can be of interest for this approach can consist of demographic data (e.g., sex or age), data on time of day, or other relevant variables such as in-/out-patient status, hospital department, or even length-of-stay.

### 3.3 Limitations

Finally, our study is prone to some limitations. Most importantly, we lack a clear healthy control group, as our data is from tertiary care only. The data encompasses some samples that come from healthy individuals (such as patients that were referred to the UMCU, but their diagnostic work-up did not confirm any diseases), but because labels are not available, we cannot identify these samples definitively. Moreover, there are some limitations considering the neighbourhood-based dimension reduction methods. One main limitation of UMAP is that the negative sampling process does not take into account the distance to the current point outside the number of nearest neighbours surrounding each point. This inaccuracy becomes more expressed if the number of samples with respect to the number of nearest neighbours is increased. The result of this is that points that are just outside the direct neighbourhood are placed incorrectly, further away in the reduced data. Additionally, UMAP is a greedy algorithm, basically requiring a copy of the original data such that incoming data can be interpolated onto the low-dimensional manifold. Furthermore, at the time of writing, neither TriMap nor PaCMAP provide a clear opportunity to embed unseen data into the space of the existing dimension reduction. This makes it harder for example to share dimension reductions between healthcare institutions, which might be beneficial, since this allows for easier interpretation of haematology measurements in the context of the overall population. Another limitation in our study is that we did not use a topology preservation metric. Scoring based on topology metrics might result in a higher ranking for the manifold approaches, as these are especially designed to preserve topology. Furthermore, we have limited our research to PCA and three manifold approaches. Of course, many more methods are available. For example, self-organising maps have been used successfully in haematological data, specifically at single-cell level [33].

## 4 Conclusion

When applying dimension reduction to high-dimensional high-volume haematology data, we found that a global statistics based reduction technique such as PCA performs systematically better than much more recent non-linear minimum-distortion dimension reduction techniques in representing the underlying data. In general, the use of dimension reduction method had limited biological performance, especially as a precursor for prediction tasks. Therefore, we advise that dimension reduction techniques are limited to data visualisation applications, e.g. for exploratory data analysis and research dissemination. The use of dimension reduction techniques as components in diagnostic pipelines may lead to decreased quality of integrated diagnostics in clinical care.

## 5 Methods

### 5.1 Descriptives

We extracted all available CBC measurements from the Abbott CELL-DYN Sapphire from 2005 to 2020 from the UPOD. We then applied rigorous quality control based on metadata retrieved from the CELL-DYN Sapphire machines, based on in-house knowledge, gained from clinical chemists and data managers. As some of the CBC measurements are only available if the sample was measured in reticulocyte mode, we imputed these missing variables using the *miceforest* package in Python, based on the Multiple Imputation with Chain Equations (MICE)[34] approach using a Random Forest approach [35]. In our data, samples were measured in reticulocyte mode by default from 2013 onwards, providing the opportunity to impute missing data before 2013, since these data could be considered Missing At Random (MAR).

Considering the possibility that extreme outliers would distort the overall quality of any dimension reduction model, we transformed white blood cell count parameters to log scale. Additionally, we decided to clip the bounds of each parameter to limit the effect of outliers, while preserving the clinical relevance of the samples. A list of the analysed variables that required clipping thresholds can be found in table S1.

### 5.2 Dimension Reduction

#### 5.2.1 Dimension Reduction Methods

One of the most frequently used dimension reduction models historically is PCA, which tries to capture data in linear combinations, using vector decomposition. It creates perpendicular components, meaning that components are not correlated to each other, and using this principle, PCA can reduce the original data into a reduced space by explaining the variance in the original data. This method is very useful when working with collinear features, as these features will be captured in the same components, since they explain the same variance in the original data. For assessing the performance of a PCA, the cumulative explained variance is often used, and this will naturally increase when the number of components are increased. PCA assumes linear relationships between variables, and assumes normally distributed variables.

Yet, as the probability exists that the original data might contain non-linear relationships, we decided to use manifold dimension reduction techniques, which are based on the theory that any space can be reduced to lower dimensions based on the shape of the data. In order to achieve this, each data point should be placed in a similar neighbourhood compared to the original space. This makes sure that local structure of the data is better preserved, i.e., that data that is similar in the original space is also similar in the reduced space. Examples of non-linear dimension reduction techniques include Uniform manifold approximation (UMAP), Triplets Manifold Approximation (TriMap), and Pairwise Controlled Manifold Approximation (PaCMAP). In addition to PCA, these methods were used in the current study to capture the large and complex CELL-DYN Sapphire dataset in lower dimension. Finally, we used Gaussian Random Projection (GRP) as a negative control. We will provide a brief overview of these techniques in this section.

Although UMAP, PacMAP and TriMap are initialised with PCA by default, the individual components of UMAP, TriMap and PaCMAP have no specific meaning, unlike PCA. For PCA, the additional explained variance diminishes when a higher number of components are used.

##### UMAP

UMAP estimates the shape of the data in the higher dimensionality using a weighted graph and then projects the graph onto the lower dimension for dimensionality reduction [1] (see figure 8). UMAP constructs a high-dimensional graph by extending branches from individual points with a radius *r* to connect the points to their neighbourhood in high-dimension. These branches then become a graph of various shapes to be projected onto the lower dimension, irrespective of distance between points. The *k*-nearest neighbours in *r* can be set, where a low *k* preserves the local structure, and a higher *k* preserves the global structure of the original data. Finally, the high-dimensional graph is projected onto a lower dimension using a force-directed graph approach, pulling together points that are close and pushing apart points are further away. This is done base on the weighted connectivity, meaning that points are drawn towards groups of points with which it has multiple connections, rather than points/clusters with singular connections. Clusters are formed based on some threshold, which also depends on the number of nearest neighbours. Increasing the *k*-nearest neighbours will result in larger groups of interconnected points, at the cost of increased computational complexity.

**Fig. 8:**
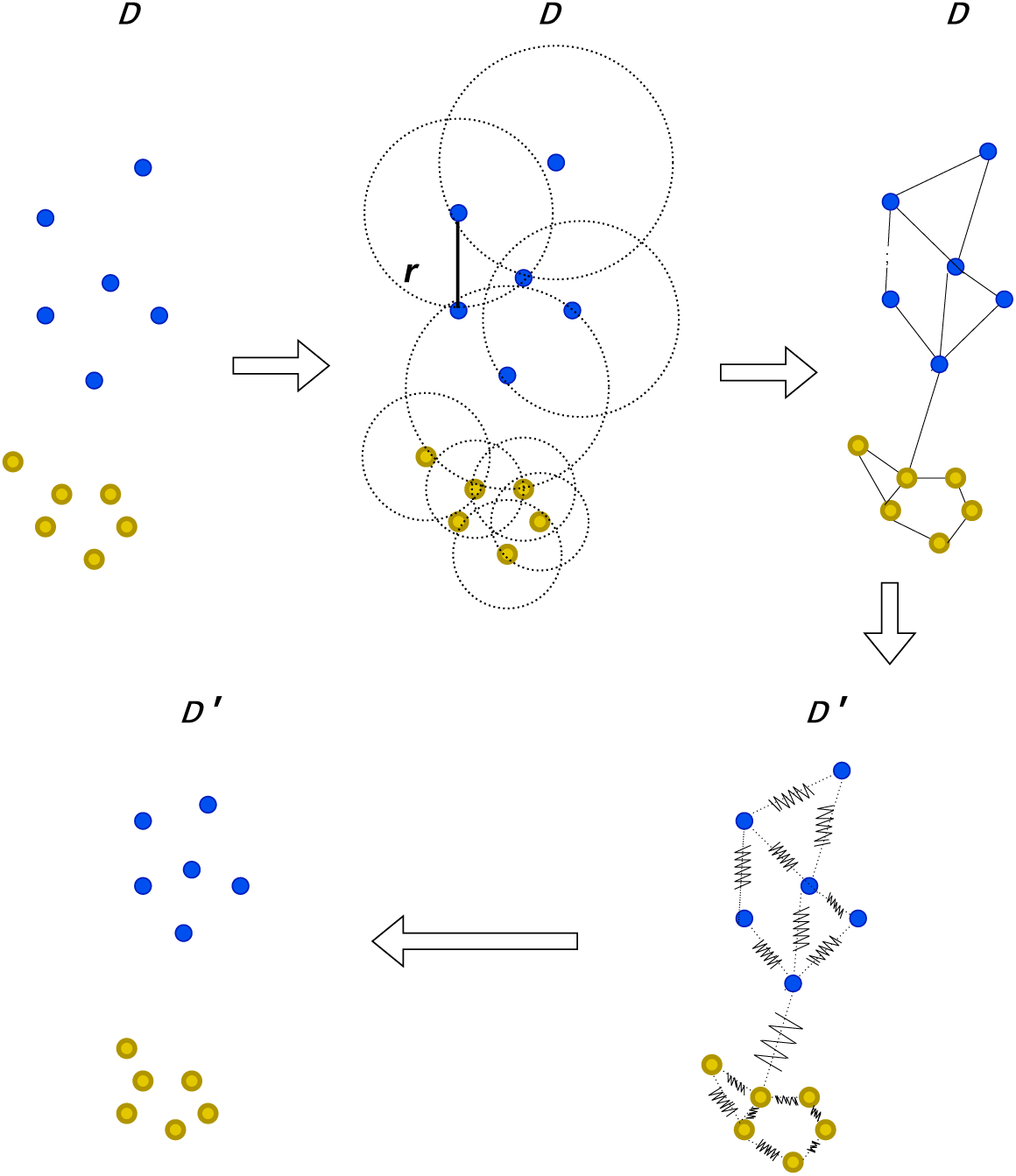
Explanation of UMAP

##### TriMap

TriMap is another manifold approach, and is primarily built around triplets constraints [3]. TriMap constructs triplets per point (*i*) and pairs this to *n inliers* (*j*) according to the distance metric used. For each of these pairings, *n outliers* are sampled (*k*) resulting in *n inliers ∗ n outliers* triplets per point (*i, j, k*). Additionally, *n random* triplets are constructed. TriMap then creates a low dimensional representation of the data where the ordering of the distances of these triplets is preserved (*d*(*i, j*) *≤ d*(*i, k*)), by weighting the triplets, according to the relative distance of *j* and *k* to *i* (figure 9).

**Fig. 9:**
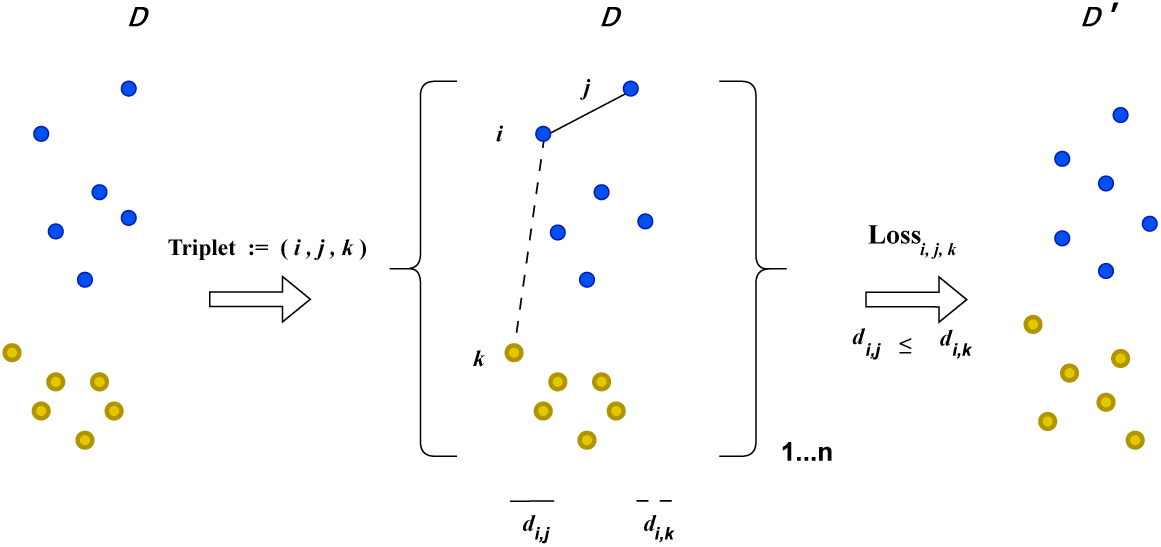
Explanation of TriMap

##### PaCMAP

Similarly to TriMap, PaCMAP samples both neighbours and non-neighbours (*Near Pairs* and *Further Pairs* respectively) in order to establish a low-dimensional representation of the original data. Contrary to TriMap, it also focusses on *Mid-Near Pairs*[2]. *Near Pairs* are the nearest neighbours based on a scaled distance metric. *Mid-near pairs* are established by sampling 6 points per observation and then selecting the second-closest point based on distance. The amount of *Mid-Near Pairs* is set by the *MN ratio*. Finally, *Further Pairs* are non-neighbours, and the amount of pairs is set using the *FP ratio*. After initializing with PCA, PaCMAP uses a weighted loss function to optimize the low dimensional representation. The loss function is primarily driven by the *Near Pairs* and *Mid-Near Pairs*, but gradually is mostly influenced by the *Near Pairs*. This means that the loss is highly increased if close points in original space are set further away in the reduced space.

##### Gaussian Random Projection

Gaussian Random Projection (GRP) is a dimension reduction technique that is based on the Johnson-Lindenstrauss lemma, which states that any high-dimensional Euclidean space can be reduced onto a lower-dimensional Euclidean space with minimal distortion (at most 1 + *ɛ*) of the pairwise distance [36], and a result by Hecht-Nielsen[37] who showed that a random selection of vectors in a high-dimensional space can be considered an orthogonal projection. Gaussian Random Projection does this by projecting original data on a randomly generated matrix with Gaussian distributions. However, the accuracy of the projection and the amount of required components for dimension reduction is highly dependent on the amount of samples and the permitted error (*ɛ*), specifically *n components ≥* 4*ln*(*n samples*)*/*(*ɛ*^2^*/*2 *− ɛ*^2^*/*3) [38]. This means that GRP can require more components than available dimensions when the number of dimensions is sufficiently low and the number of observations is high. To that end, we included GRP as a negative control for the dimension reduction quality metrics, because we would expect that this method would perform worst when dimension reduction the data to a low number of dimensions (*≤*10) because of this constraint, since our data consists of over 3 million samples.

#### 5.2.2 Parameter Tuning

We tuned the amount of neighbours used for UMAP, TriMap, PaCMAP (*n neighbours*). For UMAP and PaCMAP we were interested in the number of neighbours, but for TriMap we were interested in the number of outliers and inliers, since this is important for the construction of triplets in TriMap. Both PCA and GRP do not require any tuning on nearest neighbours, since they are not neighbours-based. Additionally, we also investigated the number of dimensions (*n components*) that were generated by all the dimension reduction methods, as this might increase the amount of information stored in the dimension reduction. For example, in PCA, the amount of total variation explained increases when the amount of components is increased. As computing numerous distinct dimension reductions and their performance is computationally expensive using a nearest-neighbours approach, we also investigated the number of samples we could use for dimension reduction purposes.

#### 5.2.3 Distance Metrics

One important step in the assessment of dimension reduction techniques is the distance metric with which we assess the distances between data points and with which we perform the dimension reduction for the manifold approaches. As mentioned above, the number of dimensions of the reduced data with Euclidean distance is dependent on the number of samples and the permitted distortion (*ɛ*). For a dataset with roughly three million samples, and roughly one hundred dimensions, this means that we are not able to project the data to a lower-dimensional Euclidean space while preserving the distortion 0 *< ɛ <* 1. This practically excludes using the Euclidean distance metric from the perspective of distance preservation, and a fractional distance metric is best suited for the description of distances in high dimensionality (*d >* 30) [39]. We decided to pursue the Manhattan distance as the simplest expression of the fractional distance.

#### 5.2.4 Dimension Reduction Quality Metrics

Two main ways that are used for dimension reduction quality metrics are evaluating the global and local structure [2]. Local structure metrics evaluate neighbourhoods of points and how well these are preserved in the reduced data, while global structure metrics evaluate how well the reduced data preserved the relationships between groups of points. In this study, both global and local distance metrics were used to find a balanced representation of the CELL-DYN Sapphire data in lower dimension. The metrics are generally rank-based, since these are insensitive to scaling. One unifying framework for rank-based metrics is the co-ranking matrix (Q-matrix) [40]. The Q-matrix compares the pairwise ranks of the original data versus the reduced data, showing the preservation of local and global distances. Calculating a Q-matrix consists of two steps. Firstly, a ranking of distances between points in both original and reduced data is calculated. Thereafter, a single matrix is constructed combining both rankings, explaining rank preservation in the low-dimensional data.

For local preservation measures, we further used the proportion of neighbouring points being preserved (the neighbourhood-kept-ratio), and the trustworthiness score. The neighbourhood-kept-ratio [6] is computed using the number of nearest neighbours **N**(*i*) for all *i* in high-dimensional space and the *k*-nearest neighbours **N***^′^*(*i*) for all *i* in low-dimensional space, where *i* is each data point. Consequently, **N**(*i*) and **N***^′^*(*i*) are compared to see the intersection between their neighbourhoods. The degree of overlap is calculated, and divided by the number of *k* to calculate a ratio for each *i*. Subsequently, this ratio is divided by the number of samples to get the average neighbourhood preservation. The trustworthiness score ranks neighbourhood points in accordance with how close they are to the observations *i* in low- and high-dimensional spaces [41]. If the ranks of neighbourhood points are misaligned in the reduced space, the metric will penalise these shifts, resulting in a lower score. A version of the trustworthiness score [41] was used in this study with help of the Q-matrix framework [42].

For global preservation measures, we used random triplet score and spearman rank correlation. The random triplet score is calculated by retrieving sets of two points (*j, k*) at random per *i* in the original data to form triplets (*i, j, k*) [2]. After this, it finds the same set of triplets in the reduced space and calculates the distance from *i* to *j* (*d_ij_*) and *k* (*d_ik_*) for both the original and the reduced data. It then orders *d_ij_* and *d_ik_* based on their distance in both datasets. The degree of order preservation indicates global structure preservation by the dimension reduction method. Five triplets per *i* were used in this study. Finally, pairwise distances can be measured using the Spearman rank correlation to assess distance preservation in the reduced data. Another strength of this method is that distance correlation is easily visualized in a graph (e.g., figure S3 and 4) to assess the correlation of distances between low- and high-dimensional spaces. To compare the different dimension reduction methods with regards to their quality metrics, we performed a the quality assessments in 10-fold, and used a T-test for comparison.

### 5.3 Preservation of biological representation

#### 5.3.1 Diurnal patterns

Because biological relevance and meaning of the data should be maintained in the dimension reduction, we assessed preservation of biological relevance from four different angles. The first and descriptive angle was by studying diurnal patterns in the reduced dataset, as the size of the original dataset allowed us to investigate large patterns within the data. We assessed the diurnal patterns in the reduced data with the use of a cosine fit, as implemented in the *CosinorPy* library[43].

#### 5.3.2 Age and sex

The second angle was to assess biological relevance by two classification tasks that should be identifiable in the data: firstly, sex prediction in samples of patients between the age of 20 to 50, as during this age-range a clear distinct difference of hemoglobin between men and women exists [44]. Secondly, prediction of samples of patients below 20 versus patients above 60 years old, as the haematological characteristics of young people are known to be distinct from older people [45]. For this purpose, we used Gradient Boosting (GB) model to capture any non-linear associations. To assess the performance of the resulting models, we decided to focus on the accuracy and the Matthews Correlation Coefficient (MCC) metric. The accuracy is the correct prediction of positive and negative cases, divided by the total amount of positives and negatives, i.e. 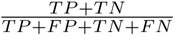, where TP, FP, TN and FN are true and false positives, and true and false negatives respectively. The MCC, or the *ϕ* coefficient, is a measure of the quality of a binary classification model that takes into account true and false positives and negatives, i.e., is a summary measure for the confusion matrix, comparable to the F1 metric. MCC is calculated as follows: 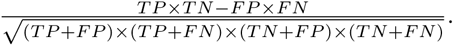 The data were analysed using 10-fold cross validation with an inner validation set (as a result of the folds) and a dedicated outer validation set. 170,000 random samples were used for training, 30,000 random samples were used for the dedicated validation set. We assessed the significance of performance change using a T-test.

#### 5.3.3 Cluster preservation

The third angle was to study the preservation of clusters of similar patients in the reduced data. We thus analyzed both the raw and the reduced data using HDBSCAN [46] and k-means clustering[47] and retrieved information on computational and analytical performances. K-means clustering retrieves a predefined number of clusters (*k*) based on the Euclidean distance towards a cluster centre, and tries to minimize the sum of distances over these *k* clusters. In practice, this can result in clusters that are of equal size and density, but are unintuitive for interpretation. HDBScan is assigning clusters based on the density of the data, and is therefore more suitable to retrieve clusters with varying densities. This increases the possibility of retrieving meaningful clusters. For this approach, we were interested in the number of clusters extracted, the Normalised Mutual Information (NMI) and Adjusted Rand Index (ARI) scores. The NMI and ARI scores are ways to report the extent of cluster preservation in the reduced data, by taking the clusters in the original data as ground truth.

#### 5.3.4 Identification of leukemia-like patients

As a final angle to assess performance of preservation of biological relevance, we investigated a specific population that is completely divergent from the general population in terms of CBC. To this end, we used samples from patients with chronic lymphatic leukemia that were diagnosed based on CBC characteristics, more specifically: very high lymphocyte counts. These samples should be clearly distinguishable in the lower dimension representation. Failure to detect these patients would significantly impact the use of the dimension reduction methods in clinical practice. To detect potentially significant differences between the populations, we used an unpaired T-test, and considered a p-value below 0.001 to be significant.

## Data Availability

The datasets generated and/or analysed during the current study are not publicly available due to privacy regulations but are available from the corresponding author on reasonable request.

https://github.com/HJJoosse/celldyn_embedder

## 6 Declarations

### Ethics approval and consent to participate

The institutional review board (Medical Research Ethics Comittee NedMec) waived the need for informed consent, as only pseudonymized data were used for a large patient sample. The study was in concordance with the declaration of Helsinki. This study was not subject to the Human Subjects Act (in Dutch: Wet Medisch-Wetenschappelijk onderzoek met mensen, WMO) and we therefore obtained a waiver for study approval from the institutional review board (Medical Research Ethics Comittee NedMec).

### Consent for publication

Not Applicable

### Authors’ contributions

Conceptualization: HJJ, SH, BvE; Methodology: HJJ, CC, BvE; Software: HJJ, CC, BvE; Validation: HJJ, AH, WWvS, SH; Formal Analysis: HJJ, CC, BvE; Investigation: HJJ, CC, BvE; Resources: SH; Data Curation: HJJ, CC, SH, BvE; Writing -original draft preparation: HJJ; Writing -review and editing: All Authors; Visualization: HJJ, CC, BvE; Supervision: AH, IEH, WWvS, SH.

## Acknowledgements

Not Applicable

