## Supplementary figures and images for "Haematology dimension reduction, a large scale application to regular care haematology data"

### Supplemental figure 1

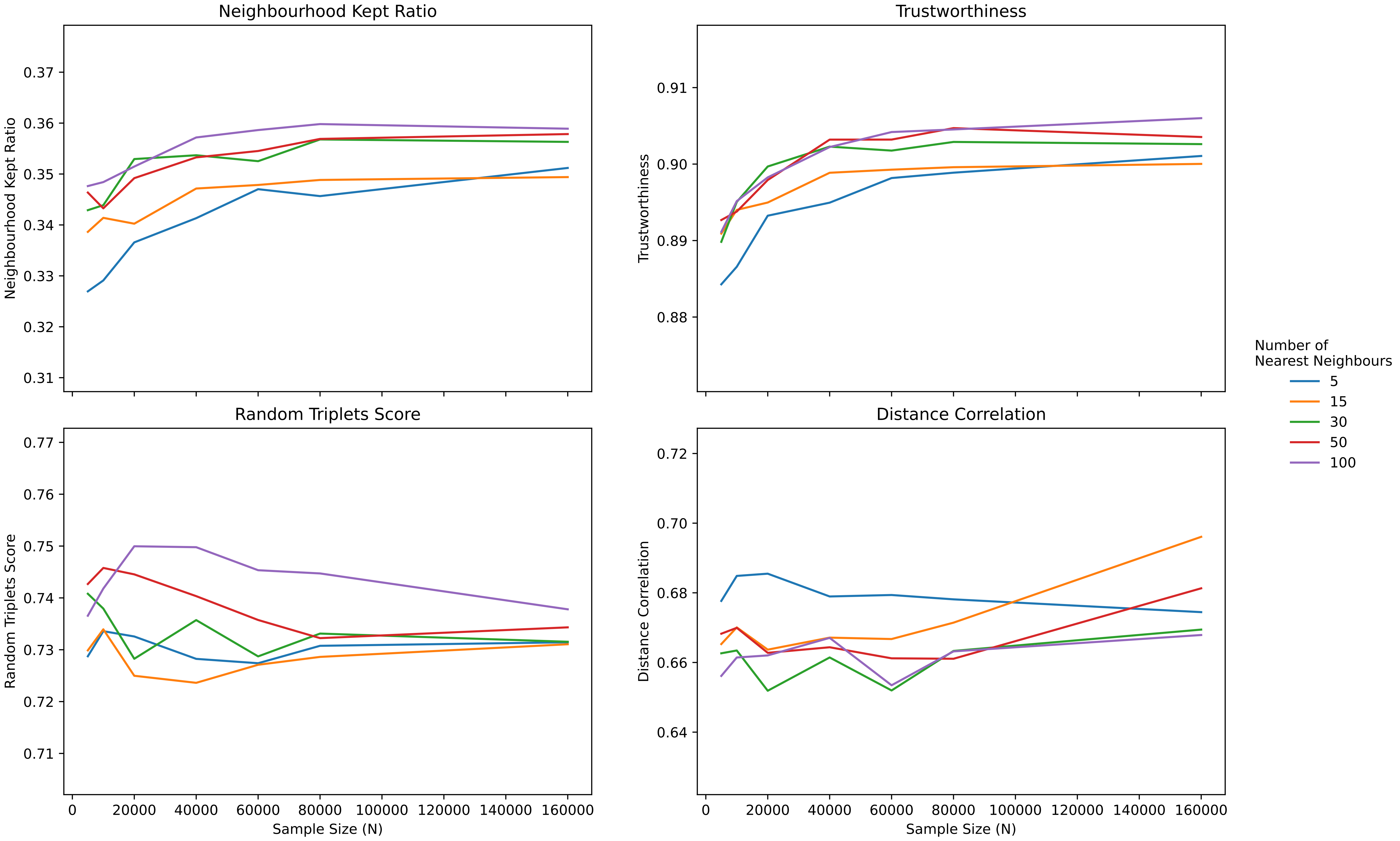

### Supplemental figure 2

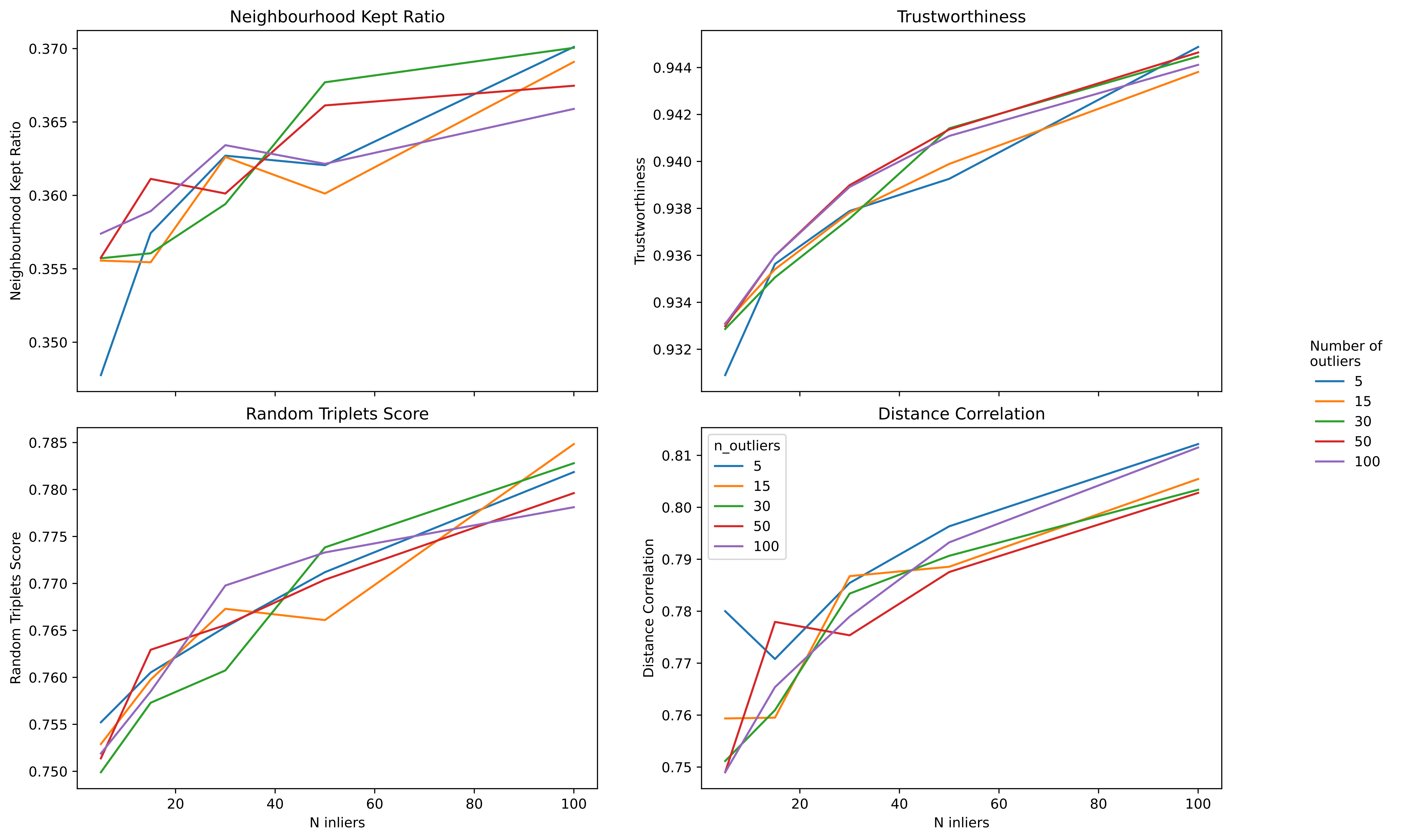

### Supplemental figure 3

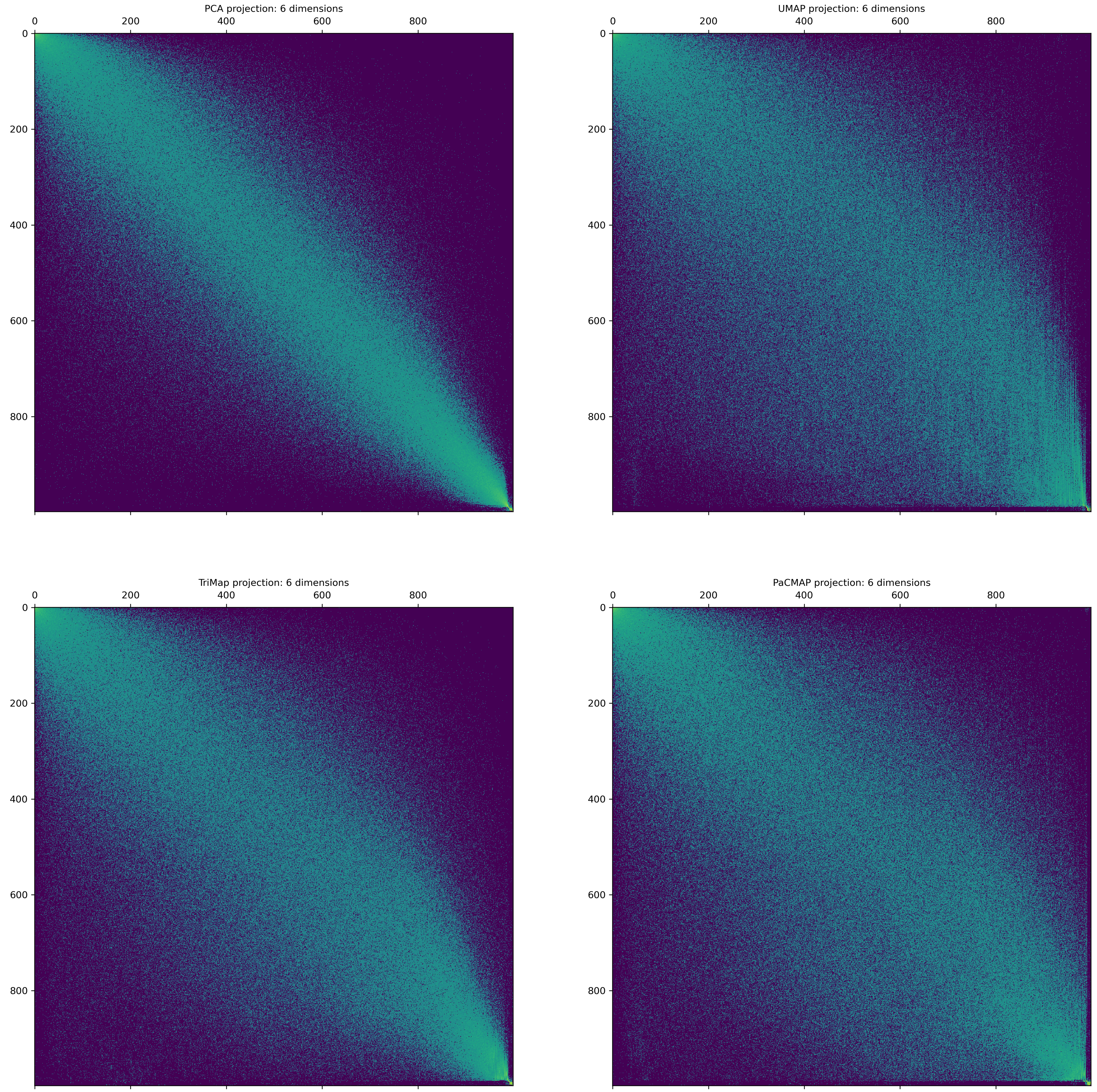

### Supplemental figure 4

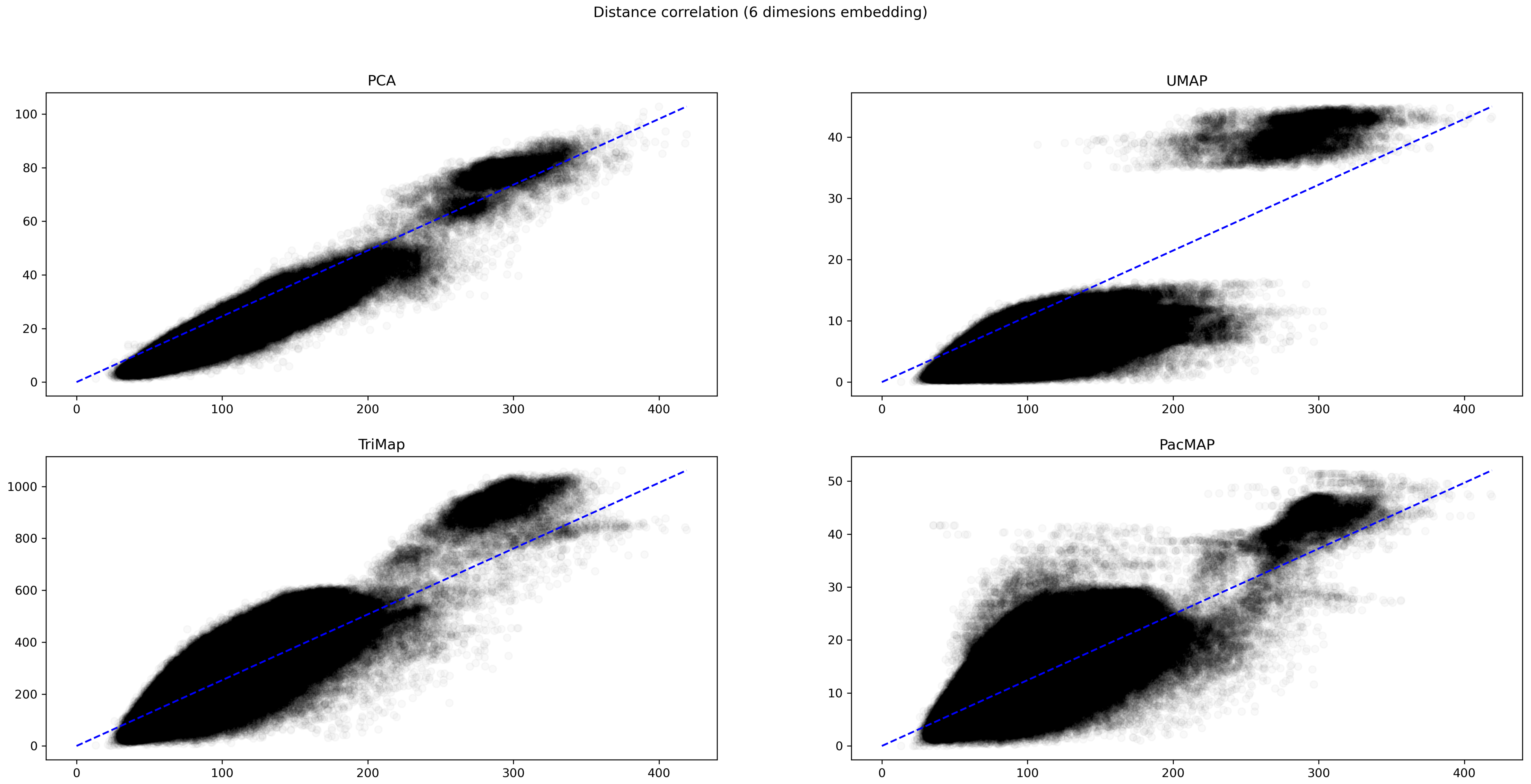

### Supplemental figure 5

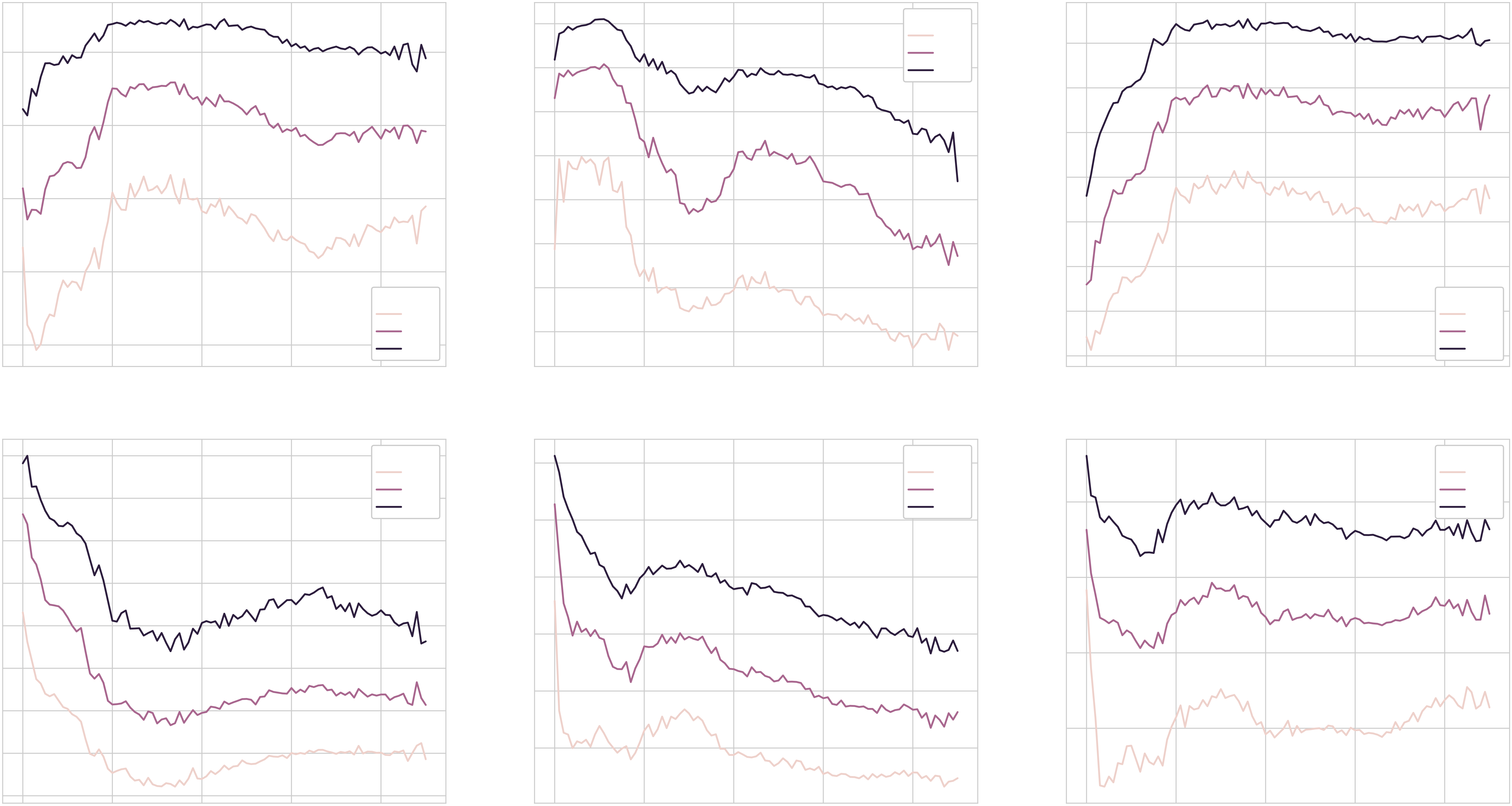

### Supplemental figure 12

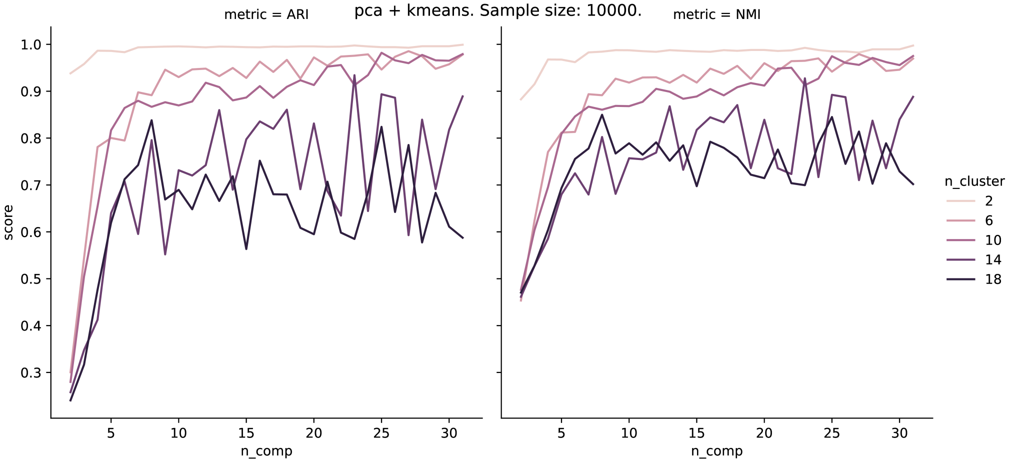

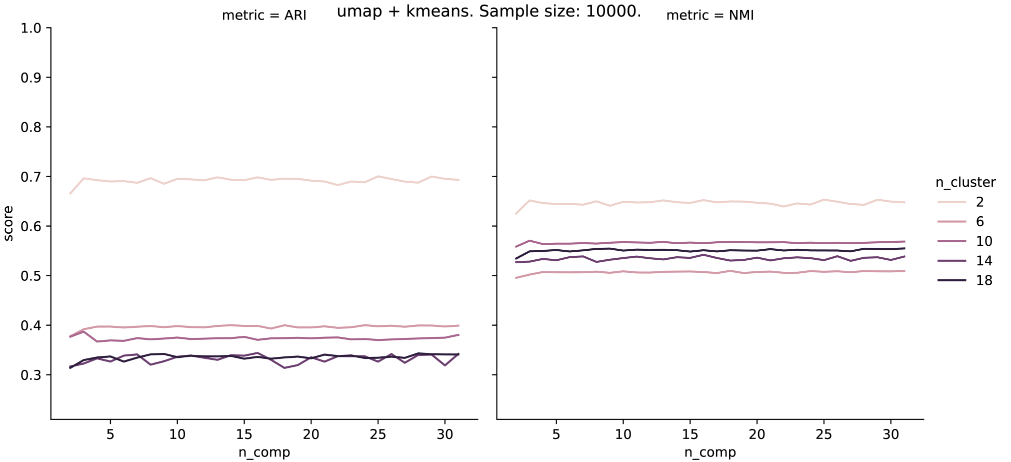

### Supplementatl figure 6

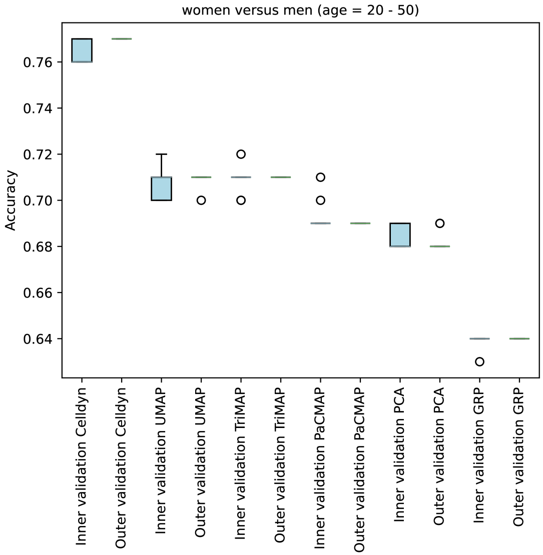

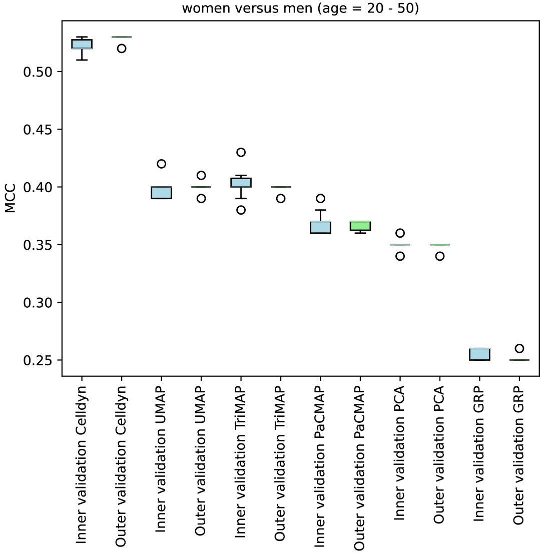
